# Asymptomatic *Genu Recurvatum* Reshapes Lower Limb Sagittal Joint and Elevation Angles During Gait at Different Speeds

**DOI:** 10.1101/2020.07.02.20144741

**Authors:** Frédéric Dierick, Céline Schreiber, Pauline Lavallée, Fabien Buisseret

## Abstract

**Purpose:** The main objective of this study is to characterize the lower limb sagittal joint and elevation angles during walking in participants with asymptomatic *genu recurvatum* and compare it with control participants without knee deformation. The secondary objective is to study the influence of walking speed on these kinematic variables.

**Methods:** The spatio-temporal parameters and kinematics of the lower limb were recorded using an optoelectronic motion capture system in 26 participants (*n* = 13 with *genu recurvatum* and *n* = 13 controls). The participants walked on an instrumented treadmill during five minutes at three different speeds: slow, medium and fast.

**Results:** Participants with *genu recurvatum* showed several significant differences with controls: a narrower step width, a greater maximum hip joint extension angle, a greater knee joint extension angle at mid stance, a lower maximum knee joint flexion angle during the swing phase, and a greater ankle joint extension angle at the end of the gait cycle. Participants with *genu recurvatum* had a greater minimum thigh elevation angle, a greater maximum foot elevation angle, and a change in the orientation of the covariance plane. Walking speed had a significant effect on nearly all lower limb joint, elevation angle, and covariance plane parameters.

**Conclusion:** Our findings show that *genu recurvatum* reshapes lower limb sagittal joint and elevation angles during walking at different speeds but preserves the covariation of elevation angles along a plane during both stance and swing phases and the rotation of this plane with increasing speed.

## 1 Introduction

Knee hyperextension or *genu recurvatum* (GR) – from Latin, *genu*: knee and *recurvare*: to bend back [34] – is an insidious condition [45] that mostly affects women [13, 14, 24] and may have negative consequence to knee joint structures [23]. It is operationally defined as a knee extension greater than 5 degrees [23]. Although there is no consensus that has set the limits of “ normality” of extension, Murphey et al. [28] suggest that the normal range of motion of the knee joint might include 10 to 15-degree extension, and a knee with more than 15-degree extension is considered as “ pathologic” [50]. GR’ s varied etiology is divided into three categories: congenital, constitutional or acquired (bone, ligament, muscle, neurological or mixed origin).

GR is typically associated with weight-bearing activities such as standing, walking, stair climbing, running, and jumping. The ground reaction force vector that acts anterior to the knee joint increases stress on the passive restraining structures that resist further knee extension [45]. Its asymptomatic (i.e. painless) form is most often bilateral, symmetrical, of constitutional “ physiological” origin related to capsuloligamentous laxity and in particular to the oblique popliteal ligament that restraint knee hyperextension [9, 26, 35]. Despite the existence of capsuloligamentous hyperlaxity, the subjects are considered “ healthy”. According to Seckin et al. [39], joint hyperlaxity in subjects with GR is observed in 11.7% (101 students out of 861) of its sample. Capsuloligamentous hyperlaxity varies with age, mainly in children, and may disappear over time [2, 42]. Depending on epidemiology, the proportion of GR can fluctuate from 10% to 25% between Eastern populations and others (African, Middle East, …) [1, 2].

Several structural changes may also be associated with GR: pseudo patella alta, excessive femoral internal rotation, genu varum or valgum, tibial varum, or excessive subtalar joint pronation [6, 23]. Passive hyperextension in isolated cadaver knee joints have shown inconsistent lesions to: the posterior joint capsule [18] and the anterior [25, 38] and posterior [18, 25] cruciate ligaments. Also, an increase contact stress on the anterior compartment of the tibiofemoral joint has been identified [25, 31] as one factor related to the increased injury and cartilage degeneration [14, 29, 30]. It can therefore be responsible for premature wear and tear of the joint cartilage and cause knee osteoarthritis. By the way, the presence of GR in patients undergoing total knee replacement surgery is not uncommon: 11.8% of the 510 knees in a recent study [40], 74.5% of which ranged from 5 to 10-degrees.

Kinematic analysis of walking is often performed to study the quality of performance of the locomotor system. The study of lower limb joint angles during walking in adult subjects with GR has already been extensively explored [5, 11, 17, 19, 20, 32]. However, these studies included neurologically impaired patients or with cruciate and collateral ligaments damage to the knee and no study has been conducted on subjects with asymptomatic GR. Furthermore, since Borghese et al. [4] proposal to measure elevation angles of the thigh, leg and foot during walking and their coordination, many studies have successfully used this methodology to better understand motor control of walking in healthy subjects [3, 8, 16] and patients with various pathologies [7, 15, 21]. To the best of our knowledge, neither lower limb joint nor elevation angles have yet been assessed in subjects with asymptomatic GR. However, the characterization of lower limb kinematics during gait in this population is important because repeated excessive movements of the knee in extension during the support phase could damage the joint structures of the knee prematurely. Moreover, since lower limb kinematics in subjects without knee deformation is strongly influenced by the speed of progression [3], it is not prohibited to hypothesize that lower limb joint and elevation angles in subjects with asymptomatic GR are speed-dependent.

The main objective of this study is to characterize the lower limb sagittal joint and elevation angles during walking in subjects with asymptomatic GR and compare it with control subjects without knee deformation. The secondary objective is to study the influence of walking speed on these kinematic variables.

## 2 Methods

### 2.1 Participants

All participants were recruited from Physical Therapy or Occupational Therapy Departments at Haute Ecole Louvain en Hainaut and from personal acquaintances. A written informed consent was signed by each participant after being informed of the experimental protocol of the study and the use of their personal data. The experimental protocol has been accepted by the local internal commission and respects the Helsinki Declaration on Ethical Principles for Medical Research Involving Human Beings. The protocol of the experiment has been approved by the Academic Bioethics Committee (B200-2018-106).

Inclusion and exclusion criteria were the following. Each participant had to be over 18 years old, a Body Mass Index (BMI) of less than 30 *kg m*^*−*2^, and intact cruciate ligaments of the knee. Integrity of the anterior cruciate ligament (ACL) of all participants was verified by two tests: the Lachman test and the anterior drawer test [10, 47], and that of posterior cruciate ligament (PCL) by the posterior drawer test [37]. Participants were excluded if they had neurologic, rheumatic, orthopedic, metabolic, genetic pathologies, were pregnant, practiced classical dance or gymnastics or intensive (competition) sport activities during the 2 weeks preceding the data acquisition. Participants with history of lower limb or spinal trauma during the 6 months preceding the data acquisition, whether or not it required physiotherapy, medical or surgical management were also excluded.

Sixty-one participants were recruited to participate in the study. Two were excluded for a BMI higher than 30 *kg m*^*−*2^. Twenty-one participants were excluded for Elhers-Danlos syndrome, ACL rupture, knee or ankle sprains, malleolar and fibular fractures. Two participants were active in competitive sports and nine declined to participate in the study (due to lack of time available). In the end, a sample of 26 participants was selected and Table 1 presents their anthropometric and clinical data. Eight participants practiced no sport (7 women, 1 man) and eighteen practiced almost one sport (11 women, 7 men). Sport activities practiced were as follows: football (4 men), volleyball (1 woman), bodybuilding (3 women, 3 men), running (4 women, 3 men), and swimming (4 women).

**Table 1.** Anthropometric and clinical data of participants for *genu recurvatum* and control groups. Knee extension is noted by a negative value. Slow, medium, and fast walking speeds adopted by the participants are given. Results of Student t-tests or Mann-Whitney tests are given for all variables. A Fisher test has been performed for the Number of GR visible in standing.

|  | *Genu recurvatum* | Control | p-value |
| --- | --- | --- | --- |
| Number of subjects (men/women) | 13 (9/4) | 13 (9/4) | – |
| Age (years) | 21.2±1.7 | 20.5 1.3 | 0.257 |
| Height (cm) | 167.9±0.1 | 168.2±0.1 | 0.916 |
| Body mass (kg) | 65.4±11 | 63.3±11.0 | 0.521 |
| Body Mass Index (kg m<sup>-2</sup>) | 23.2±3.4 | 22.3±3.1 | 0.505 |
| Passive knee extension amplitude (degrees) | -12±3 | -3±1 | <0.001 |
| Active knee extension amplitude (degrees) | -8±3 | -1±1 | <0.001 |
| Number of GR visible in standing (yes/no) | 8/5 | 0/13 | <0.001 |
| Slow walking speed, S1 (m s<sup>-1</sup>) | 0.68±0.02 | 0.68±0.02 | 0.814 |
| Medium walking speed, S2 (m s<sup>-1</sup>) | 1.33±0.03 | 1.33±0.03 | 0.627 |
| Fast walking speed, S3 (m s<sup>-1</sup>) | 1.78±0.04 | 1.78±0.04 | 0.638 |

The sample was divided into two groups: GR (*n* = 13, knee extension higher than 5 degrees) or control (*n* = 13, knee extension lower than or equal to 5 degrees), according to the manual passive maximum knee extension angle value (see Table 1: Passive knee extension amplitude) measured in supine position. Maximum knee extension angle value was also assessed while the participant contracted the quadriceps (see Table 1: Active knee extension amplitude). For the passive measurement, the experimenter’ s (P.L.) hand holds were identical to those proposed by Ramesh et al. [36], with the proximal hand placed on the anterior part of the knee and the distal hand below the heel. The experimenter extended the knee by lifting the leg from the heel [36]. For the active measurement, a 10-cm bolster was placed under the distal tibia [41, 42] and the participant was asked to extend the knee as fully as possible. The measurement of knee extension angulation was performed using a long arm goniometer as the angle formed between a line from the lateral femoral epicondyle to the greater trochanter and a line from the lateral femoral epicondyle to the lateral malleolus. A negative value indicated knee hyperextension. No attempt was made to assess the amplitude of knee extension in the standing position as proposed by several authors [22, 24, 48] since measurement technique in supine position is more reliable, as the participants are better able to maximally extend the knee in this position [42]. GR in standing position was visually observed by the experimenter without telling it to the participants (see Table 1: Number of GR visible in standing).

### 2.2 Experimental protocol and data acquisition

Kinematic data were collected by the same experienced operator using a motion capture system (Vicon Motion Systems Ltd, Oxford Metrics, Oxford, United Kingdom) consisting of 8 optoelectronic cameras (Vero v.2.2) with a sampling frequency of 120 Hz. Sixteen passive reflective markers with a size of 14 mm in diameter were used. The placement of these markers, using double-sided adhesive tape, was applied according to the lower body Plug-in-Gait model (Oxford Metrics, Oxford, United Kingdom). The model includes two marker positions; twelve markers symmetrically placed on anatomical landmarks identifiable by palpation, i.e., on the anterior and posterior superior iliac spines (ASI and PSI), on the knee flexion/extension axis (KNE), on the lateral malleolus along the imaginary line passing through the trans-malleolar axis (ANK), on the head of the 2^nd^ metatarsal (TOE) and on the calcaneus (HEE). The asymmetry of the other four markers, namely those positioned at the lower third of the lateral side of the thigh (THI) and leg (TIB), was necessary for easily differentiate between the left and right side. Participants were asked to wear only shorts for men and a bra for women in order to maximize the visualization of the markers and their appropriate positioning throughout the gait measurements.

The motion capture system was calibrated according to the standard procedure. A static recording was first made for a few seconds in the typical position, corresponding to the standing position, with the knees slightly bent with the shoulders raised to 90 degrees and the elbows extended. Participant’ s anthropometric data, namely height, weight, length of lower limbs and width of knees and ankles were collected. Dynamic data collection was realized while the participants were walking, wearing running shoes, on a motorized treadmill. All subjects performed three walking trials of 5-minutes at slow (S1, around 2.5 km h^*−*1^), medium (S2, around 5 km *h*^*−*1^), and fast (S3, around 6.5 km *h*^*−*1^) speeds. The sequence of walking speeds was randomized for each participant. S1, S2, and S3 were normalized for each participant using the Froude (*Fr*) number [49], and defined as *Fr* = *v*^2^*/gL*, where *v* is the walking speed (m s^*−*1^), *g* is the acceleration due to gravity (9.81 m s^*−*2^), and *L* is the length of the lower limb, measured here as the vertical distance (m) between the greater trochanter of the femur and the ground. *Fr* values were 0.055, 0.21, and 0.37, respectively for S1, S2, and S3. Mean and SD values of S1, S2, and S3 adopted by the participants are shown in Table 1. Although they were all familiar with treadmill walking, the participants were given a 4-minutes period to familiarize with the treadmill and acquisition environment at each of the imposed speeds. Only the last minute of walking was recorded by the motion capture system.

After the recording of the walking trials, Vicon Nexus software (v.2.7.1, Oxford Metrics, Oxford, UK) was used for reconstruction and 3D modelling of markers, leading to the trajectories **x**_*a*_ = (*x*_*a*_, *y*_*a*_, *z*_*a*_) of the markers *a* in a cartesian frame were *z* is the vertical axis, *x* is antero-posterior axis, and *y* is the medio-lateral axis. A Plug-in Gait Dynamic pipeline was applied with a Woltring quintic spline algorithm with a mean square error value of 10. The data were then exported with the ASCII standard in a csv file for further processing. This file included all the 3D positions of the markers as well as the value of the hip, knee, and ankle angles as function of time. Only the angles in the sagittal plane were retained for further analysis. The gait cycles have all been normalized from 0% (heel strike) to 100% (the following heel strike). Identification of heel strike events was realized in Nexus software that computes the duration (*s*) of each of the gait cycles. Data analysis was performed over 15 consecutive gait cycles for each speed. Maximum angle of hip joint extension (*H*_1_) and flexion (*H*_2_), knee joint angle value at heel strike (*K*_1_), maximum angle of knee joint flexion during the stance phase (*K*_2_) and extension at mid stance (*K*_3_), maximum angle of knee joint flexion during the swing phase (*K*_4_), maximum angle of ankle joint flexion after heel strike (*A*_1_), ankle joint angle at foot flat (*A*_2_), maximum angle of ankle joint extension during the stance phase (*A*_3_), and ankle joint angle at the end of the gait cycle (*A*_4_) were collected with R Studio software (v.3.4.4, Boston, MA, United States of America) using homemade functions. All parameters computed are presented in Figure 1. In participants with GR, the analysis was limited to the side with the greatest mean *K*_3_-value for medium speed (left knee for all but one women, right knee for all men). Arbitrarily, the left knee was selected for all the control subjects.

**Fig. 1.**
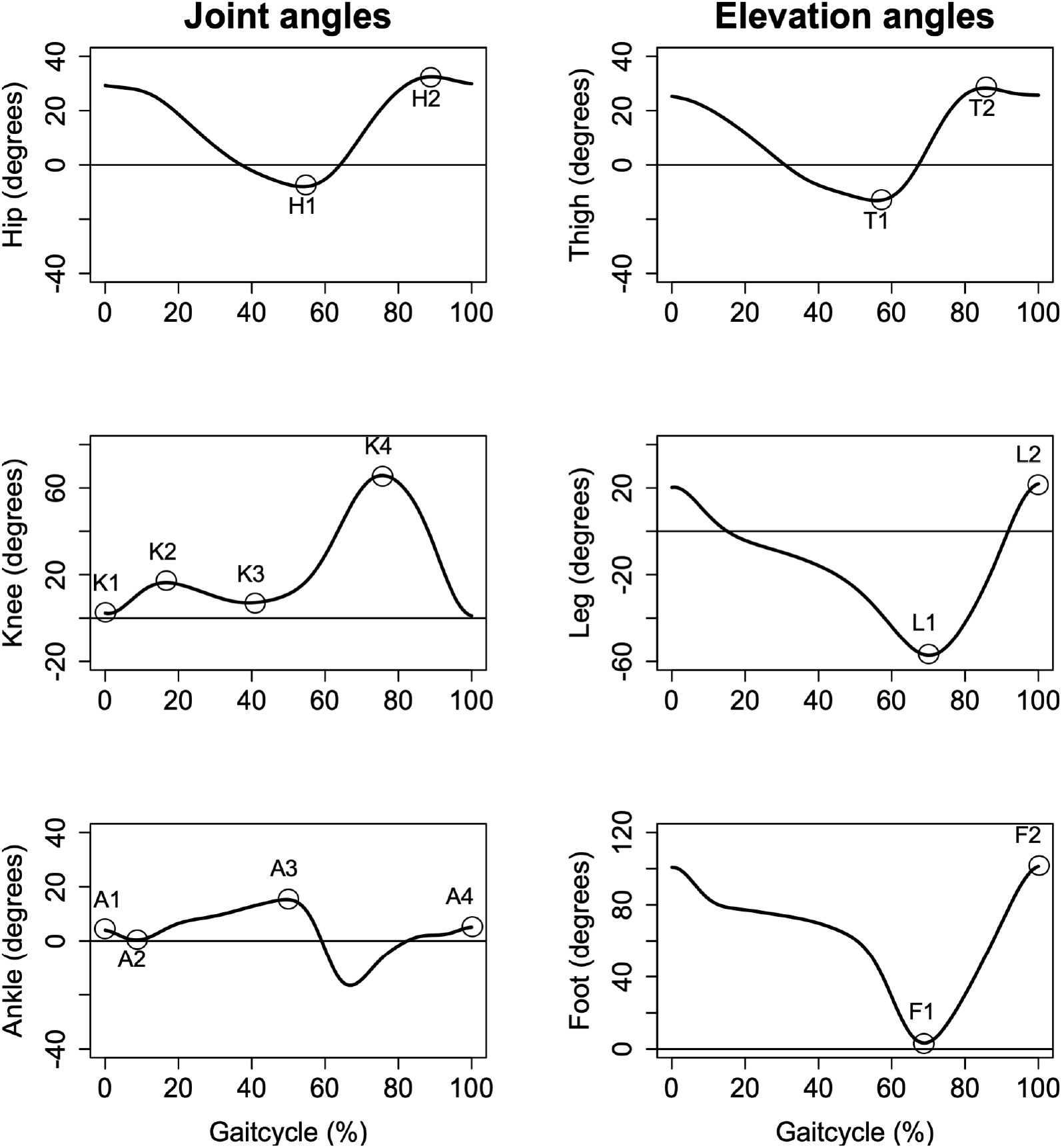
Parameters collected for lower limb joint and elevation angles analyses.

Vertical ground reaction forces were collected by an instrumented treadmill (N-Mill, Motekforce Link, The Netherlands) at a sampling frequency of 500 Hz and spatio-temporal parameters of gait were automatically determined using the manufacturer’ s software (CueFors 2, Motekforce Link, The Netherlands), including step and stride lengths (Step L and Stride L, m), step width (Step W, m), step rate (Step R, steps min^*−*1^), gait cycle duration (GCD, s), and unipodal stance duration (USD, s).

Walking kinematics was also assessed by computing lower limb elevation angles during the gait cycle in the sagittal plane [4]. The elevation angle *α*, for a given segment *i* (thigh, leg or foot), was calculated according to the equation: 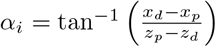, where *p* and *d* subscripts correspond respectively to the proximal and distal points of the segment. Minimum and maximum elevation angles for thigh (*T*_1_, *T*_2_), leg (*L*_1_, *L*_2_), and foot (*F*_1_, *F*_2_) segments were collected using R Studio.

Inter-segmental coordination between thigh, leg, and foot angles of elevation was assessed by computing the covariance plane [4]. A regression on the curve made by these three angles, measured over the 15 gait cycles, allows to find the cartesian equation of the covariance plane, hence the vector **U** = (*U*_*foot*_, *U*_*leg*_, *U*_*thigh*_) normal to the plane. The indices refer to the axis related to a given angle.

### 2.3 Statistical analysis

Means and standard deviations (SD) of all parameters were computed. The statistical analyses were performed using SigmaPlot software (v.11.0, Systat Software, San Jose, CA, United States of America). A two-way repeated measures (RM) analysis of variance (ANOVA) was realized to assess the influence of status (GR or control) and walking speeds (S1, S2, S3) on all variables. The interaction of these two parameters (Status*×* Speed) was also tested. *Post-hoc* Holm Sidak tests were performed when RM ANOVA was significant. One-sample t-test was performed on the angle of deviation between the plane of the GR participants and the plane of the controls. The significance threshold for all statistical tests was set at 5%.

## 3 Results

Anthropometric data and walking speeds were not significantly different between GR and control groups (Table 1). The maximum passive and active knee extension amplitudes were significantly different between the groups, the GR participants showing higher values (Table 1). Visual observation of GR in standing position was confirmed in 8 participants on 13 in the GR group and was never observed in the control group (Table 1).

Table 2 presents the results of the spatio-temporal parameters. No significant differences were observed between the groups (Status), except for Step W. GR participants walked with a narrower step width compared to controls (Table 2). Walking speed had a major significant effect on all spatio-temporal parameters, except for Step W. Step R, Step L, and Stride L increased significantly with speed, and GCD and USD decreased significantly with speed (Table 2). No significant Status *×* Speed interaction was observed for spatiotemporal parameters.

**Table 2.** Two-way RM ANOVA results for spatio-temporal parameters, lower limb joint and elevation angle parameters in participants at slow, medium, and fast walking speeds, dor *genu recurvatum* (GR) and control groups. Parameters displayed are: Step rate (Step R), gait cycle duration (GCD), unipodal stance duration (USD), step and stride lengths (Step L and Stride L), step width (Step W), maximum angle of hip joint extension (*H*_1_) and flexion (*H*_2_), knee joint angle value at heel strike (*K*_1_), maximum angle of knee joint flexion during the stance phase (*K*_2_) and extension at mid stance (*K*_3_), maximum angle of knee joint flexion during the swing phase (*K*_4_), maximum angle of ankle joint flexion after heel strike (*A*_1_), ankle joint angle at foot flat (*A*_2_), maximum angle of ankle joint extension during the stance phase (*A*_3_), ankle joint angle at the end of the gait cycle (*A*_4_), minimum and maximum elevation angles for thigh (*T*_1_, *T*_2_), leg (*L*_1_, *L*_2_), and foot (*F*_1_, *F*_2_). The vector normal to the covariance plane, **U**, is finally shown.

|  | Slow speed (S1) |  | Medium speed (S2) |  | Fast speed (S3) |  | p-value |  |  |
| --- | --- | --- | --- | --- | --- | --- | --- | --- | --- |
|  | GR | Control | GR | Control | GR | Control | Status | Speed | St×Sp |
| Step R (*steps min*<sup>-1</sup>) | 78.20±5.75 | 80.92±5.23 | 108.80±5.13 | 110.77±3.72 | 125.31±6.05 | 127.85±5.37 | 0.211 | <**0.001** | 0.874 |
| GCD (s) | 1.55±0.15 | 1.51±0.11 | 1.11±0.06 | 1.09±0.05 | 0.97±0.04 | 0.95±0.04 | 0.441 | <**0.001** | 0.625 |
| USD (s) | 1.03±0.08 | 1.00±0.07 | 0.70±0.04 | 0.69±0.03 | 0.60±0.03 | 0.59±0.03 | 0.223 | <**0.001** | 0.384 |
| Step L (m) | 0.54±0.04 | 0.52±0.05 | 0.74±0.04 | 0.72±0.04 | 0.86±0.06 | 0.83±0.05 | 0.175 | <**0.001** | 0.833 |
| Stride L (m) | 1.07±0.08 | 1.02±0.07 | 1.48±0.08 | 1.44±0.06 | 1.66±0.20 | 1.66±0.10 | 0.403 | <**0.001** | 0.459 |
| Step W (m) | 0.10±0.03 | 0.12±0.04 | 0.09±0.03 | 0.12±0.01 | 0.10±0.02 | 0.12±0.02 | **0.010** | 0.841 | 0.903 |
| $H_1$ (degrees) | -13.2±6.3 | -8.1±4.0 | -18.3±5.7 | -13.9±4.4 | -20.5±5.0 | -15.9±4.5 | **0.019** | <**0.001** | 0.738 |
| $H_2$ (degrees) | 25.6±7.1 | 28.6±4.0 | 30.5±6.2 | 32.8±4.4 | 36.0±7.2 | 37.6±1.3 | 0.284 | <**0.001** | 0.539 |
| $K_1$ (degrees) | -2.2±5.0 | 1.3±3.3 | -3.0±5.7 | -0.8±2.9 | -1.8±5.1 | 0.3±3.5 | 0.111 | 0.051 | 0.437 |
| $K_2$ (degrees) | 7.8±3.1 | 8.9±3.7 | 13.1±5.2 | 15.2±2.9 | 19.0±5.3 | 20.1±3.3 | 0.288 | <**0.001** | 0.754 |
| $K_3$ (degrees) | -2.8±4.0 | 1.8±1.9 | -4.2±4.8 | 1.8±3.1 | -17.7±4.4 | 0.5±3.0 | <**0.001** | <**0.001** | **0.010** |
| $K_4$ (degrees) | 53.5±4.4 | 54.9±3.6 | 59.5±5.8 | 63.1±3.5 | 59.1±3.6 | 63.2±2.9 | **0.038** | <**0.001** | 0.123 |
| $A_1$ (degrees) | 1.9±3.4 | -0.8±3.2 | 3.1±3.8 | -0.2±2.9 | 3.8±3.8 | 2.6±3.2 | 0.065 | <**0.001** | 0.062 |
| $A_2$ (degrees) | 17.3±2.8 | 15.8±3.3 | 15.9±3.7 | 14.8±2.1 | 13.5±3.6 | 12.6±3.0 | 0.271 | <**0.001** | 0.807 |
| $A_3$ (degrees) | -4.8±6.4 | -4.8±6.1 | -11.9±6.3 | -16.2±6.4 | -16.8±5.0 | -17.6±5.6 | 0.393 | <**0.001** | 0.130 |
| $A_4$ (degrees) | 10.3±2.6 | 6.3±2.7 | 9.8±2.9 | 5.1±3.0 | 12.0±3.1 | 8.7±3.3 | **0.010** | **0.010** | 0.459 |
| $T_1$ (degrees) | -21.3±5.0 | -17.8±3.7 | -25.7±3.8 | -21.5±3.9 | -29.7±4.0 | -25.1±3.6 | **0.009** | <**0.001** | 0.510 |
| $T_2$ (degrees) | 17.7±3.9 | 19.7±4.4 | 22.2±3.2 | 24.1±4.3 | 25.1±4.0 | 26.0±4.0 | 0.329 | <**0.001** | 0.241 |
| $L_1$ (degrees) | -48.2±4.4 | -47.9±2.4 | -54.5±2.4 | -55.6±2.3 | -58.8±3.4 | -58.9±1.2 | 0.744 | <**0.001** | 0.325 |
| $L_2$ (degrees) | 16.6±4.8 | 13.9±2.4 | 22.1±3.5 | 21.1±1.8 | 23.4±3.1 | 22.9±2.4 | 0.201 | <**0.001** | 0.137 |
| $F_1$ (degrees) | 22.4±9.2 | 22.5±6.6 | 7.0±5.3 | 3.3±5.3 | 0.5±3.8 | -0.8±3.9 | 0.301 | <**0.001** | 0.443 |
| $F_2$ (degrees) | 98.1±5.3 | 92.2±4.2 | 105.9±2.8 | 101.4±2.9 | 110.0±2.7 | 106.0±2.6 | <**0.001** | <**0.001** | 0.405 |
| $U_{foot}$ | 0.75±0.04 | 0.73±0.03 | 0.79±0.02 | 0.78±0.01 | 0.80±0.02 | 0.80±0.01 | 0.539 | <**0.001** | 0.389 |
| $U_{leg}$ | -0.63±0.05 | -0.63±0.04 | -0.59±0.02 | -0.58±0.02 | -0.58±0.01 | -0.58±0.02 | 0.531 | <**0.001** | 0.596 |
| $U_{thigh}$ | -0.20±0.06 | -0.25±0.05 | -0.19±0.05 | -0.23±0.05 | -0.12±0.08 | -0.14±0.05 | **0.040** | <**0.001** | 0.508 |

St×Sp: Status×Speed

Figure 2 and Table 2 present the results for the lower limb joint angle parameters. GR participants had a significant greater maximum hip joint extension angle (*H*_1_), and a greater maximum knee joint extension angle at mid stance (*K*_3_) than controls. Maximum knee joint flexion angle during the swing phase (*K*_4_) was significantly lower in GR participants. Ankle joint extension angle at the end of the gait cycle (*A*_4_) was significantly greater in GR participants. Walking speed had a major significant effect on all lower limb joint angle parameters (Table 2), except for knee joint angle value at heel strike (*K*_1_). The only significant Status*×*Speed interaction was observed for maximum angle of knee joint extension at mid stance (*K*_3_).

**Fig. 2.**
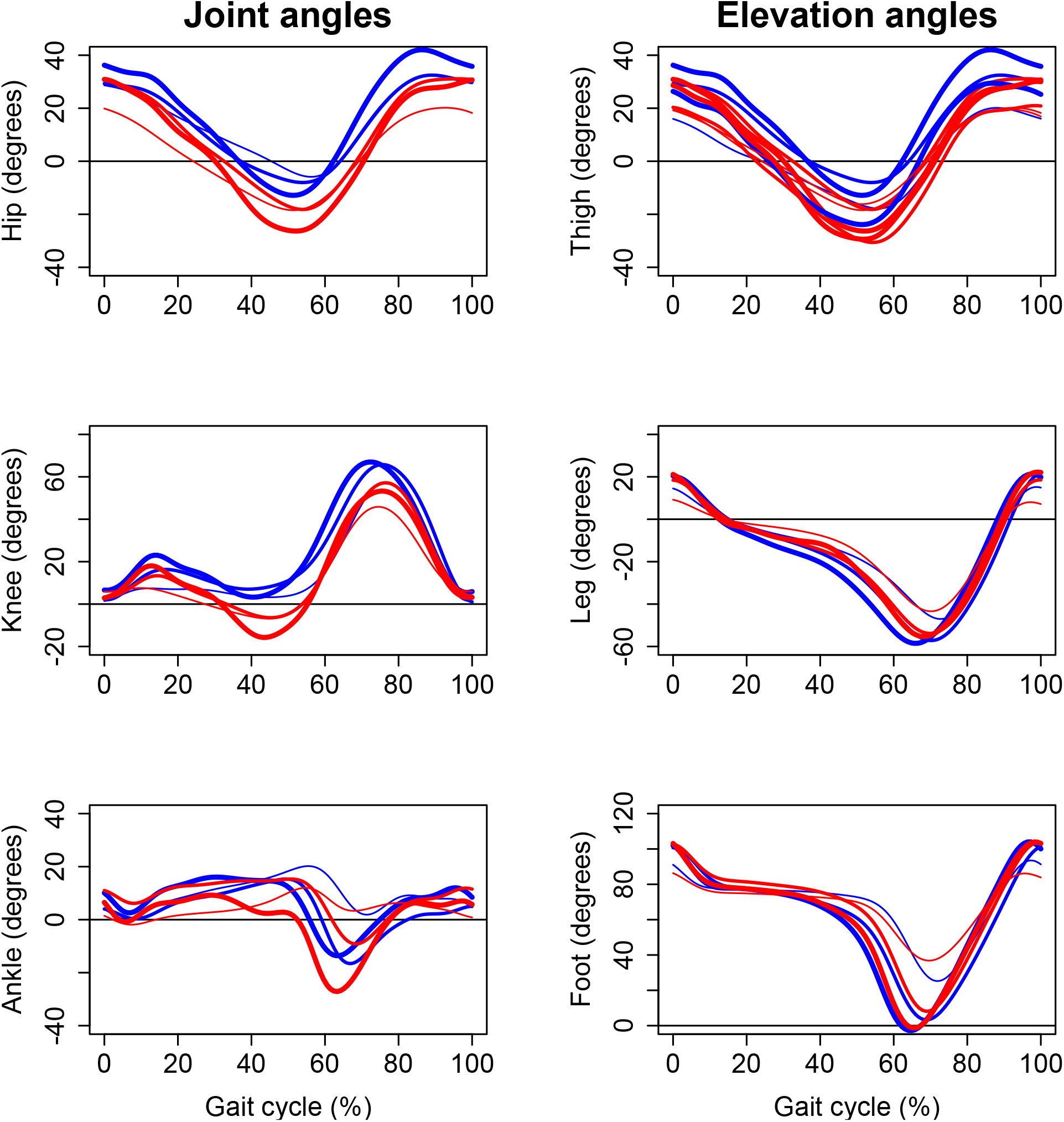
Lower limb joint (hip, knee and ankle) and elevation (thigh, leg, foot) angles shown for one GR participant (red curves) and one control participant (blue curves). Time is normalized in % of gait cycle. The thickness (thin, medium, thick) of the curves correspond to the 3 speeds (slow, medium, fast).

Two lower limb elevation angle parameters were significantly different between the groups. Participants with GR had a greater minimum thigh elevation angle (*T*_1_) and a greater maximum foot elevation angle (*F*_2_) compared to controls (Table 2). Walking speed had a significant effect on all lower limb elevation angle parameters (Table 2). No significant Status *×* Speed interaction was observed for lower limb elevation angles parameters.

Table 2 also presents the results for the covariation planes. Lower limb planar covariation was preserved in participants with GR. It is readily checked from inspection of foot *versus* leg elevation angles (Fig. 3). Only *U*_*thigh*_ shows a significant difference between the groups (Table 2). Figure 3 show 3D plots of thigh, leg, and foot elevation angles for one GR participant and control at slow, medium and fast speeds. These plots draw regular gait loops constraint to a planar plane whose orientation rotates significantly about the long axis of the loop with status and increasing speed (see *U*_*thigh*_ results in Table 2). Walking speed had also a significant effect on *U*_*foot*_ and *U*_*leg*_. *U*_*thigh*_, *U*_*leg*_, and *U*_*foot*_ results for GR and control participants as function of walking speed are presented in Fig. 4. Note that no significant Status *×* Speed interaction was observed for covariance plane parameters (Table 2).

**Fig. 3.**
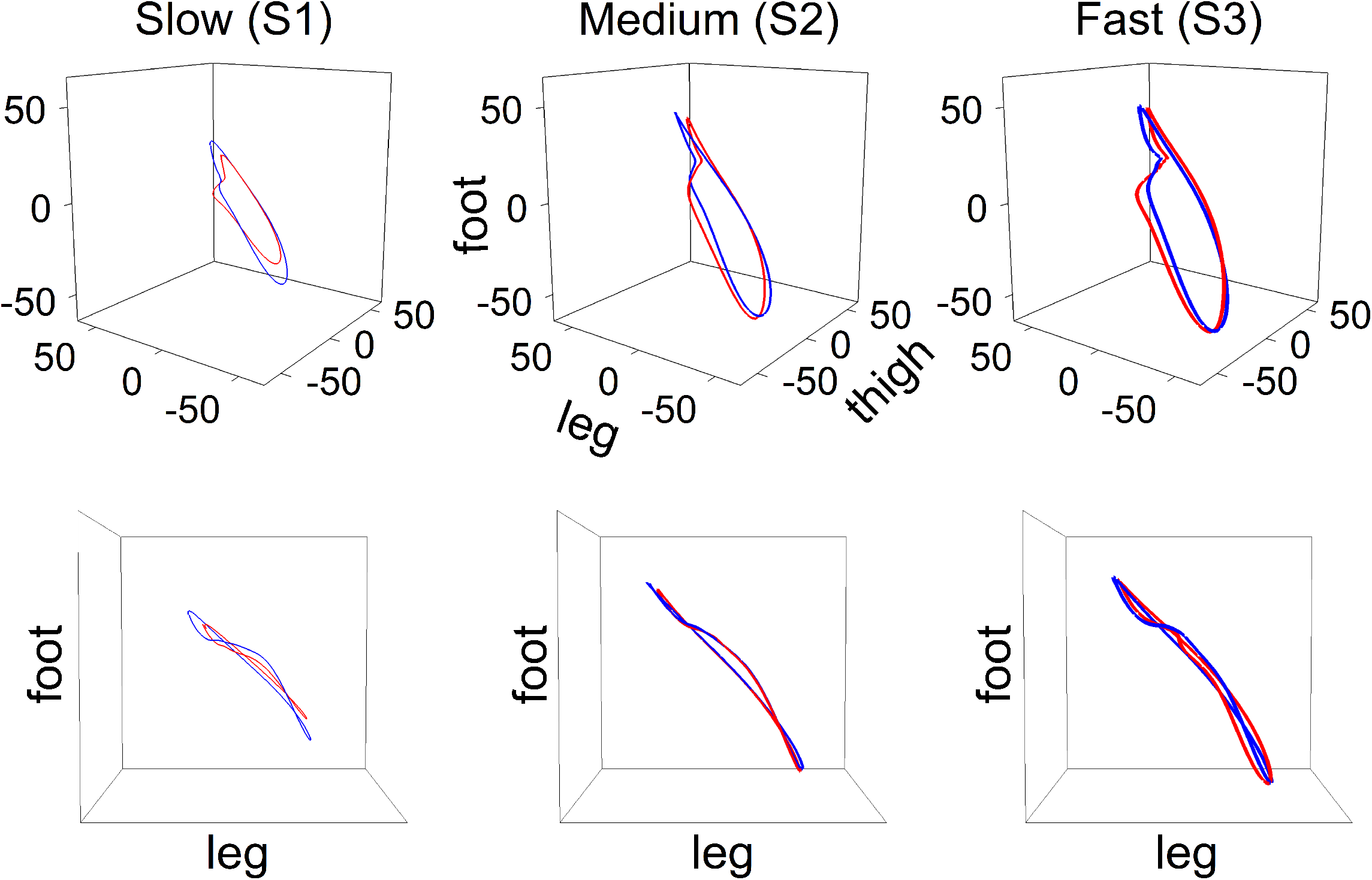
3D plots of thigh, leg, and foot elevation angles for one GR participant (red curves) and one control participant (blue curves) at slow (left panel), medium (central panel) and fast speeds (right panel). The convention for the thickness of the curves is the same as in Figure 2.

**Fig. 4.**
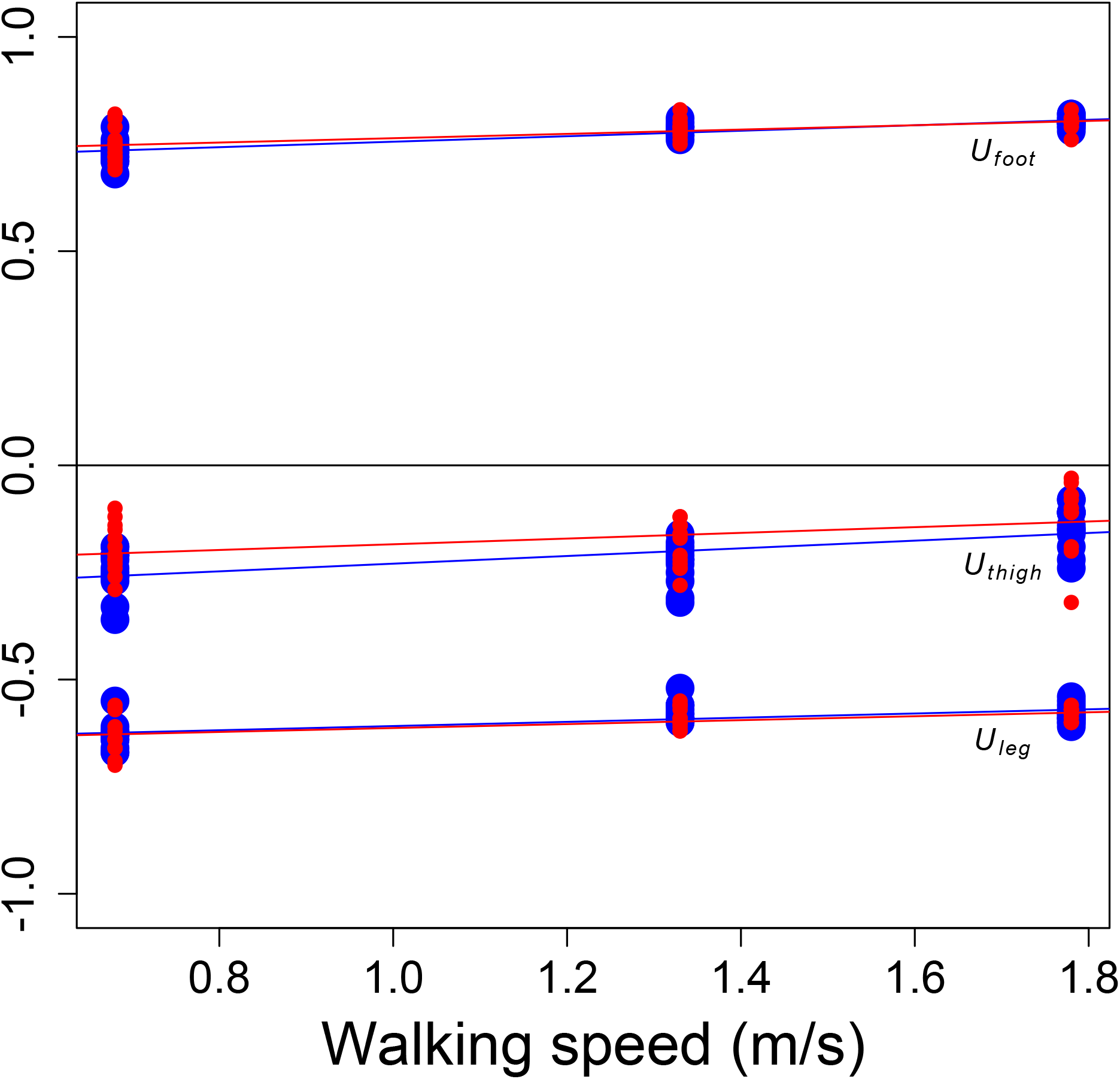
*U*_*thigh*_, *U*_*leg*_, and *U*_*foot*_ values for GR (red points) and control (blue points) participants as function of walking speed. Least-square linear regressions are also shown.

## 4 Discussion

To the best of our knowledge, it is the first study that characterizes lower limb kinematic changes in asymptomatic individuals with GR compared to individuals without knee deformation and assesses the influence of walking speed. Characterization of kinematic changes during gait in asymptomatic individuals with GR is very important since repeated abnormal movements of the knee could lead to premature degeneration of its anatomical structures. In addition to classical joint angles assessment, elevation angles and their planar covariation were also computed. The latter methodology is more and more frequently used in gait analysis of healthy individuals but also in clinical gait analysis for patients with neurological or orthopedic impairments [7, 15, 21]. Their relevant nature makes them an essential tool for researchers who wish to fully characterize the kinematics of walking.

The operational definition of GR, with a manually measured knee joint extension angle greater than 5 degrees [22, 23] was used to divide our participants in GR and control groups. This definition was also adopted in several previous studies in asymptomatic subjects [45, 46]. However, we regret the lack of consensus about the position and modalities that must be adopted to assess GR. Here, we observed a magnitude of passive knee hyperextension of 12*±* 3 degrees in GR group, slightly higher than that observed by Teran-Yengle et al. [46] (9.2 *±* 2.5 degrees). This difference could be explained by the difference in measurement procedure. In Teran-Yengle et al. [46], the ankle was resting on a 10-cm support during the measurement, while our experimenter extended the knee by lifting the leg from the heel, as proposed by Ramesh et al. [36].

At the clinical examination, we observed a significant difference of passive knee hyperextension between our groups. The magnitude of -3*±* 1 degrees observed in controls is different than the value reported in normative data for female subjects aged between 20 and 44 years old (1.6 *±* 2.8 degrees) [43]. In their study, the sample size is larger and even though the supine position was adopted for the measurement of passive knee extension, no precision regarding the position of the experimenter’ s hands to apply the forces is mentioned and could explain the observed difference.

During gait at intermediate speed, the magnitude of knee hyperextension at midstance in GR group (-4.2*±* 4.8 degrees) is smaller than that previously reported in other studies with a similar population (8.7*±* 3.3 degrees in Teran-Yengle et al. [45] and 7.4*±* 2.4 degrees in Teran-Yengle et al. [46]). However, these studies only explored a walking speed of 1.3 m s^*−*1^, making comparison of our results at slow and fast speeds impossible. Note that at fast walking speed, S3 (1.78 m s^*−*1^), we observed an important knee hyperextension of - 17.7*±* 4.4 degrees in GR group. At fast speed, the knee hyperextension observed in GR was more important to the one observed in clinical passive examination. Therefore, in future studies including young asymptomatic participants with GR, we recommend to also assess them at faster walking speeds, specifically when attempting to correct knee hyperextension [45, 46]. Additionally, Seo et al. [40] stated that GR deformation could be responsible for premature wear of the joint cartilage and therefore induce knee osteoarthritis. Since at fast speed, the knee hyperextension observed is important, damages could be more important than at medium and slow speeds. As a preventive measure, women with GR should avoid walking at high speeds in order to limit the risk of premature wear to the knee joint structures.

No differences between the two groups were observed for the spatio-temporal variables, except for the step width. GR participants adopted a narrower step width than controls. This finding could be explained by the excessive knee adduction moment associated to hyperextension movement [32].

Maximum knee hyperextension angle was mainly marked during the stance phase of gait, *K*_3_, compared to heel strike, *K*_1_. Our findings therefore diverge from those of Teran-Yengle et al. [45] and Teran-Yengle et al. [46] who observed maximum knee extension at heel strike in an asymptomatic population but are in agreement with those of Noyes et al. [32] in symptomatic participants. At the end of the stance phase, near toe-off, the flexion action of the gastrocnemius muscles typically counterbalances the anterior ground reaction force that is attempting to move the knee into hyperextension [45]. It is therefore not forbidden to hypothesize that an altered electrical activity of these muscles could be responsible for the GR pattern observed. This hypothesis must be explored in future studies since, to our knowledge, no study recorded electrical activity of gastrocnemius muscles during gait in asymptomatic GR participants. Maximum knee joint flexion angle during the swing phase, *K*_4_, is lower in GR than controls. GR participants actually show a knee joint angle pattern that is shifted downward with respect to control participants. It appears to be a signature of GR gait pattern.

The gait is a complex sequence of movements which includes both open and closed kinematic chains existing during the swing and stance phases, respectively [44]. In an open kinematic chain, the knee joint angle can be adjusted without incurring any changes in more proximal/distal joints but in a closed kinematic chain, both ends of the chain are fixed and any adjustment in the knee joint angle could reciprocally results in altered angles in more proximal/distal joints. By the way, complex interactions between trunk, hip and ankle during the stance phase of walking were previously hypothesized in subjects with GR of neurological origin [19]. Here, we believe that it is also the case in GR of orthopedic origin since significant hip/thigh kinematic modifications at the end of the stance phase (*H*_1_, *T*_1_) were observed compared to controls. A significant hip hyperextension characterized the GR participants. In addition to the GR itself, this hip hyperextension at the end of the stance phase could be achieved by: hyperextending the ankle or hyperextending the lumbar spine. Our findings reveal that it would be mainly related to hip/thigh kinematic modifications (*H*_1_ and *T*_1_ values were significantly different between groups), and therefore probably to lumbar spine position, and not hyperextension of the ankle (*A*_3_ values were not significantly different between groups). Unfortunately, the kinematics of the spine has not been recorded and this hypothesis remains to be verified. In the current state of knowledge, hip hyperextension seems to be specific to GR of orthopedic origin since it is a major difference compared to subjects with a neurological GR, such as after a stroke, who show reduced maximum hip extension during the stance phase [27, 33].

In addition to the observations made in the closed kinematic chain, modifications in the open kinematic chain were highlighted. Ankle/foot modifications in GR group were observed at the end of the swing phase (*A*_4_, *F*_2_) compared to controls. Maximum ankle extension and maximum foot elevation angles were greater in GR. Even if these observations complete the previous studies that only explored knee kinematics in asymptomatic GR participants [45, 46], we do not believe that these differences are of major importance since they cannot be involved in degenerative processes.

In 3D plots of lower limb elevation angles, we observed regular gait loops constrained in a plane as it was also observed in previous studies [3, 4] and constitute the first kinematic law. As also observed by these researchers, the covariance plane rotates about the long axis of the loops with increasing walking speed (*U*_*foot*_, *U*_*leg*_, and *U*_*thigh*_) and this constitute the second kinematic law. Here, we can conclude that these two kinematic laws observed in healthy participants [3] are preserved in case of GR.

Some limitations must be acknowledged. First, global joint mobility assessment using Beighton’ s score [2] was not realized and would have identified participant’ s hypermobility. Second, it would have been interesting to take into account the menstrual cycle phase of the participants. However, evidence of an association between hormonal fluctuations and knee joint ligaments laxity is low [12].

## 5 Conclusions

The assessment of lower limb joint and elevation angles at slow, medium, and fast walking speeds constitutes a strong point of the study. Our findings show that walking speed profoundly influences joint/elevation angles in the two groups and that assessment of GR at fast speed is required to complete the understanding of the kinematics in this asymptomatic population. All parameters computed were significantly different between the two groups, except knee joint angle value at heel strike, *K*_1_ that was very close to be significant. Despite the asymptomatic nature of GR deformity, our kinematic findings complete those obtained in previous studies by showing the existence lower limb kinematic modifications in closed and open chains. The hip hyperextension observed during the stance phase seems to be specific to GR of orthopedic origin and is of paramount importance for the implementation of verbal instructions [32] or real-time biofeedback [45, 46] provided during rehabilitation protocols attempting to correct knee hyperextension of orthopedic origin during gait.

## Data Availability

Data are available on request.

## Acknowledgements

With the financial support of the European Regional Development Fund (Interreg FWVl NOMADe). The authors thank François Lavallée for technical support during data analysis.

## List of abbreviations

*A*_1_: maximum angle of ankle joint flexion after heel strike
*A*_2_: ankle joint angle at foot flat
*A*_3_: maximum angle of ankle joint extension during the stance phase
*A*_4_: ankle joint angle at the end of the gait cycle
ACL: anterior cruciate ligament
ANOVA: analysis of variance
BMI: body mass index
*F*_1_: minimum elevation angle for foot
*F*_2_: maximum elevation angle for foot
*Fr*: Froude number
GCD: gait cycle duration
GR: *genu recurvatum*
*H*_1_: maximum angle of hip joint extension
*H*_2_: maximum angle of hip joint flexion
*K*_1_: knee joint angle at heel strike
*K*_2_: maximum angle of knee joint flexion during the stance phase
*K*_3_: maximum angle of knee joint extension at mid stance
*K*_4_: maximum angle of knee joint flexion during the swing phase
*L*_1_: minimum elevation angle for leg
*L*_2_: maximum elevation angle for leg
PCL: posterior cruciate ligament
RM: repeated measures
S1: slow walking speed
S2: medium walking speed
S3: fast walking speed
Step L: step length
Step R: step rate
Step W: step width
Stride L: stride length
*T*_1_: minimum elevation angle for thigh
*T*_2_: maximum elevation angle for thigh
*U*: vector normal to the covariance plane
*U*_*foot*_: foot component of **U**
*U*_*leg*_: leg component of **U**
*U*_*thigh*_: thigh component of **U**
USD: unipodal stance duration

